# AI Segmentation of Vestibular Schwannomas with Radiomic Analysis and Clinical Correlates

**DOI:** 10.1101/2023.06.15.23291439

**Authors:** Mikhail Milchenko, Kevin Cross, Harrison Smith, Pamela LaMontagne, Satrajit Chakrabarty, Kaamya Varagur, Rano Chatterjee, Patel Bhuvic, Albert Kim, Daniel Marcus

**Author notes:** Corresponding Author Info: Mikhail Milchenko.

## Abstract

Vestibular schwannoma (VS) is a benign, slow growing tumor that may affect hearing and balance. It accounts for 7-8% of all primary brain tumors. Gamma knife radiosurgery (GKRS) is a common treatment option for VS. Magnetic resonance imaging (MRI) is employed for diagnosis, surgery planning, and follow-up of VS. Long-term follow-up determines efficacy of VS treatment. Identifying MRI-derived markers to improve management of VS is challenging. This study describes MRI processing pipeline that automatically segments VS and investigates stability and outcome predictive power of radiomic MRI features.

We first preprocessed and segmented available pre-GKRS T1-weighted post-contrast MRI images in VS patients, using a Convolutional Neural Network (CNN) developed on DeepMedic framework. Then, we compared CNN and manual segmentations, extracted radiomic features from both manual and CNN segmentations of VS, and, finally, evaluated robustness of extracted features and clinical outcome analyses based thereof.

We found that homogeneity, robust maximum intensity and sphericity were the most robust across segmentations. We also found that maximum and minimum intensities were most predictive of tumor growth across all segmentation methods and subject cohorts. We used retrospective post-GK SRS data collected in our institution to build the processing pipeline for unsupervised segmenting of VS. This pipeline is released as a Docker image integrated with XNAT (extensible neuroimaging archive toolkit), an established open research imaging database platform^15^. Generated segmentations can be viewed and edited in the XNAT-based online OHIF (Open Health Imaging Foundation) viewer^16^ in real time.

## 1. Introduction

The optimal management of asymptomatic vestibular schwannomas (VS), benign tumors arising from the sheath of the eighth cranial nerve, is uncertain. Overall, VS affect 1.5 per 100,000 per year^1^. 95% are sporadic, while the remainder are associated with inherited cancer predisposition syndromes, e.g., the autosomal dominant syndrome Neurofibromatosis Type 2. In the current MRI era, the diagnosis of VS is being made earlier, while identified tumors are, on average, smaller and often asymptomatic^2^. In most cases, an observational approach is first taken^2^. Often, if growth or symptoms occur, active treatment, including surgical resection or radiation approaches such as stereotactic radiosurgery (SRS) or fractionated radiation therapy is pursued.

Several studies have attempted to describe the natural history of VS^3,4^. In a 2020 study from Denmark, where 40 years of national VS data and outcomes were prospectively collected, 1075 intrameatal and 1237 extrameatal tumors were initially observed^3^, while 20 intrameatal and 1272 extrameatal tumors were initially treated. Of 774 initially-observed extrameatal tumors for which growth data were available, 156 (20%) went on to grow > 2 mm. Of those, 91% grew fewer than 10 mm. In the extrameatal observational cohort, 22% went on to surgery and 4% went on to radiation. Marinelli et al analyzed growth in patients undergoing observation using volumetric analysis (defined as >20% growth) and found among 952 patients that 80% of tumors grew at 5 years, reflecting greater sensitivity of volumetric analysis. Walsh et al analyzed the growth of conservatively managed tumors over a 12-year period in Toronto and found that 45% of initially observed tumors grew > 1 mm, 37% were stagnant or grew < 1 mm, and 17% of tumors spontaneously decreased in size. Failure of initial conservative management occurred in 15% of patients.

Currently, radiomic and biological markers are under investigation to predict which tumors will grow and require therapy and which will remain at a similar size or contract. Serum and CSF biomarkers, while able to interrogate directly the molecular pathways implicated in tumor growth, are limited by their reliance on phlebotomy or lumbar puncture and for the most part have not been adopted clinically^5^. Radiomics, which applies quantitative analysis to clinical imaging data, offers a non-invasive and orthogonal means of tumor analysis but has also not been widely adopted clinically^6^. Since 2012, several studies have attempted to identify radiomic features prognostic of subsequent VS growth, with varying success. Truong et al^7^ did not find correlation between MRI texture analysis and growth in 78 patient cohort; Yang et al^8^ found that high tumor texture inhomogeneity produced better response to gamma knife SRS (GK SRS) in 336 patients; Langenhuizen et al.^9^ (61 patients) and D’Amico et al.^10^ (38 patients) used various texture features to predict pseudoprogression and tumor growth.

There remains great promise in radiomics as accessibility grows. However, the ability to perform radiomics depends on an accurate and reproducible volumetric mask of a tumor. In the above studies, all tumors were manually delineated. As multi-institutional imaging databases such as The Cancer Imaging Archive (TCIA) expand and the amount of available imaging data grows exponentially, there will be increasing demand for streamlined processes to segment tumors automatically for radiomic assessment, and for correlation of radiomic features with cellular and molecular features.

Several groups have conducted trials of AI-guided volumetric segmentation of VS imaged with contrast MRI^11–14^, with study size ranging between 89 and 324 subjects, and reporting Dice coefficient between the manual segmentation and convolutional neural network (CNN) output between 85 and 92%.

Given the current advances in segmentation techniques and radiomic analyses, the next question that must be investigated is the effect of intrinsic variability in segmentation output using different methods, on the subsequent analyses that involve radiomic features. We therefore designed our present study to first preprocess and segment available pre-GK SRS contrast MRI images of VS patients, with one of the state-of-the-art CNN approaches. Then, we compare the CNN output to manual segmentations, extract basic radiomic features from both manual and CNN segmentations of VS, and, finally, evaluate robustness of extracted features and clinical outcome analyses based on extracted features. We found that homogeneity, robust maximum intensity and sphericity were the most robust across segmentations. We also found that maximum and minimum intensities were most predictive of tumor growth across all segmentation methods and subject cohorts. We used retrospective post-GK SRS data collected in our institution to build the full processing pipeline capable of segmenting VS in an automated fashion. The resulting pipeline is released as a Docker image that can be run as standalone or built into XNAT (extensible neuroimaging archive toolkit), an established open research imaging database platform^15^, where it is integrated into the online OHIF (Open Health Imaging Foundation) viewer^16^ with ability to generate and edit VS segmentations in real time.

## 2. Methods

### 2.1. Study participants, MR imaging and lesion outlines

For this study, we identified a retrospective cohort of 229 participants who underwent stereotactic radiosurgery between 2001 and 2018. Patients had to be 18yrs or older and have a new or suspected diagnosis of unilateral VS. Patients who have had prior surgery or radiation therapy for the VS or suspected meningioma diagnosis were excluded. Each included VS was treated with single-fraction Gamma Knife radiosurgery (GK SRS) with the Leksell Gamma Knife (Elekta AB, Stockholm, Sweden). T1c images were acquired on five different scanner models manufactured by Siemens, as well as one GE Genesis Signa scanner, with 1.5T field strength, 1 mm median in plane resolution, 1.5 mm median slice thickness, 29 ms median repetition time, 8 ms median echo time, and 40° median flip angle.

After preprocessing, 158 out of 229 participants (79 female, median age 58.9, age range [23.1 – 91.3]) passed our image registration quality control and were used for all further analyses (Figure 1). Available lesion outlines were extracted from the planning workstation and converted to lesion masks for this study labeled as “GK” (Figure 1) for 95 of the original 158 participants. Two groups were created for those with (N = 95, 49 female, median age 58.5, age range [27.2-87.3]) and without (N = 63, 30 female, median age 59.8, age range [23.1-91.3]) GK SRS lesion masks.

**Figure 1.**
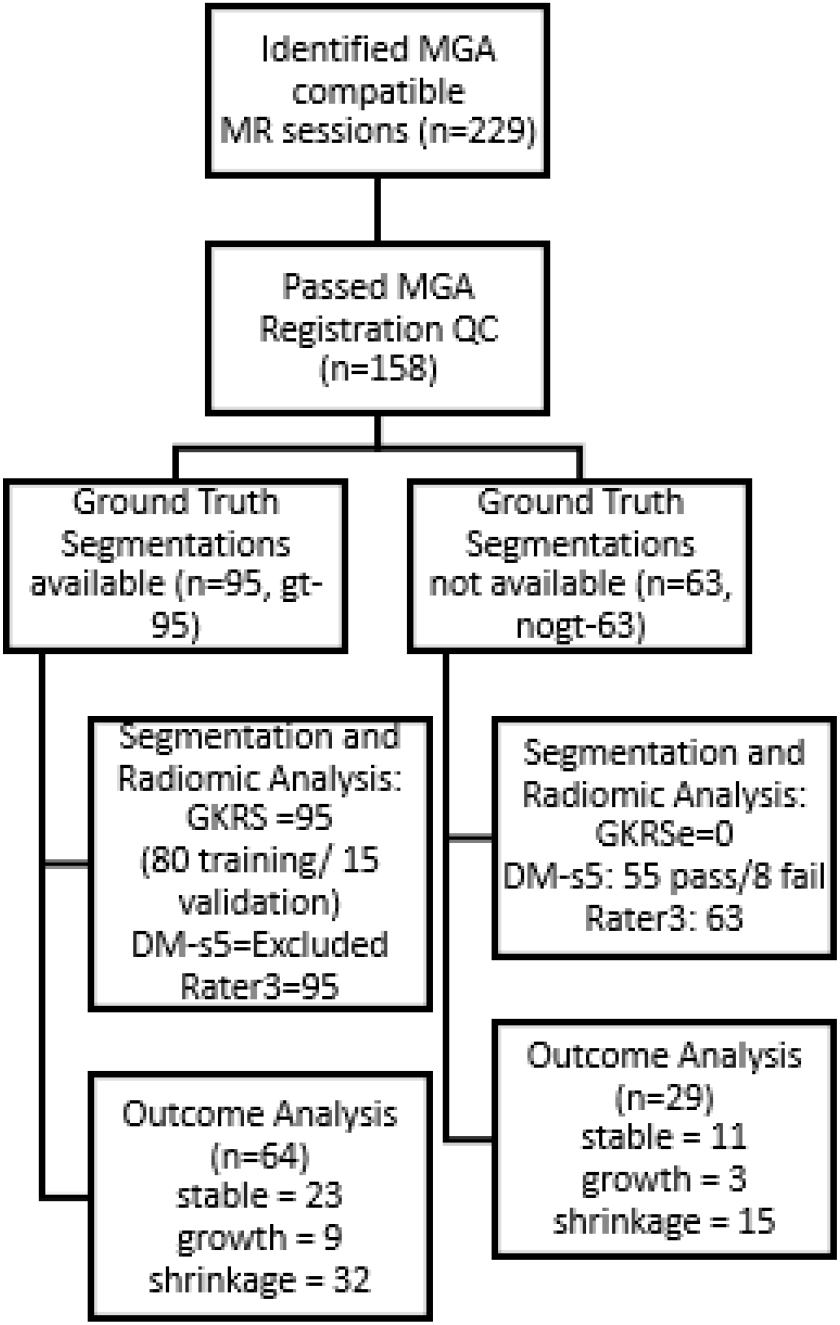
Size and structure of datasets in the study.

Out of the 158 that passed preprocessing, 93 cases had follow-up imaging and were used for outcome analyses. Follow-up outcomes were tracked until a scan showed tumor growth as reported by a neuroradiologist or until the last available scan. Imaging was acquired typically every 6 months for first year and then every year thereafter, with follow-up time average of 56 months and range of 6-96 months. The retrospective study and data sharing were approved by the Washington University in St. Louis Institutional Review Board.

### 2.2. Preprocessing of T1c

#### 2.2.1. Initial preprocessing

Since VS is well localized in the brain, we proceed by spatially coregistering all the T1c images to a reference space using multimodal glioma analysis (MGA) pipeline^17^. As a spatial reference, we use a locally developed variant of average T1-weighted template image. This template, referred to as Tal111, is linearly aligned to Talairach space^18^ and features 1 mm isotropic voxels and 176x208x176 mm field of view in axial orientation with anterior-posterior commissure trajectory coinciding with the Y axis. This brain atlas image space is also linked via 12-parameter Affine transform to the widely used MNI152 atlas^19^. Following MGA completion, we inspected corresponding registration quality control (QC) reports to identify failed registrations.

After MGA was run for the first time on all cases, we found that 67% of the T1c acquisitions had partial brain coverage between 10% and 60%. Since registration algorithms commonly expect full brain images to be available to drive registration, this led to 153 out of 229 cases failing spatial alignment to reference space, out of which 126 had partial brain field of view (FOV) coverage.

To register the acquisitions where only a fraction of brain was imaged, we applied an iterative procedure where cases that were successfully aligned at the initial stage, drove the registration of failed cases (Figure 2). Specifically, we selected 29 brain images with partial FOV that passed registration QC and had the lesion masks available, resampled them to the standard template space and created an average template image referred further as pFOV-29. This template was then binarized to create a partial brain mask. We also averaged the Affine transform matrix from each of these brain images to standard space, to obtain an average transform. Thus obtained mask and average transform were used to initialize the registration using the locally developed registration tool^20,21^. We used this tool to optimize the Affine transform first using 6 DOF (rigid body), and then 12 DOF optimization. For objective function that quantifies the alignment between two images, we used conjugate metric gradient measure^20^. This registration procedure was applied to all originally failed cases with both partial and full brain coverage. As a result, 158 T1c brain images were correctly registered and used in subsequent analyses (Figure 1). Out of the 71 remaining failed cases, 59 had partial brain coverage.

**Figure 2.**
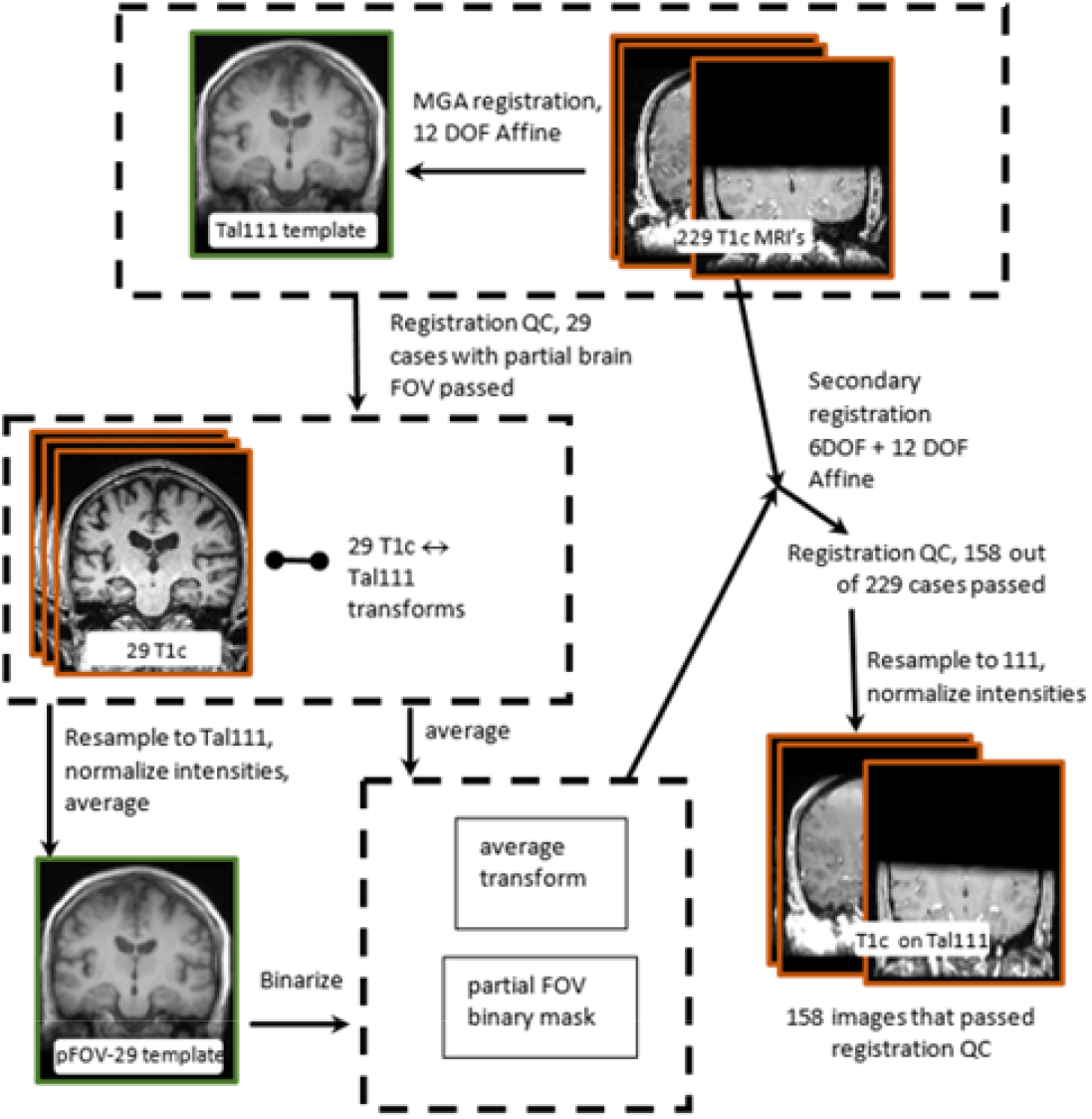
Iterative registration scheme of T1c MRI with partial brain field of view.

Following registration which estimated the 12-parameter Affine transform to Tal111 reference space, all T1c images and available lesion label masks were resampled to this space. For resampling of T1c images and masks, we used trilinear and nearest neighbor interpolation, respectively. Using standard reference space for all brains provided key anatomical prior information to the subsequent automated segmentation procedure.

The final pre-processing step comprised two variants of normalization, linear and piecewise linear. For linear normalization, we used z transform. In particular, we computed mean and standard deviation within the pre-defined brain region in Tal111 space, and linearly transform image intensities so that mean and standard deviation of brain signal become 0 and 1, respectively. In piecewise linear normalization, we utilized the multi-tissue standardization of intensities (STI) approach^22^ where modes of joint histograms between image and standard template are matched to construct a piecewise linear intensity mapping. Because clinical T1c images often lack contrast between white and grey matter (WM and GM), we modeled both of these as a single class (WM+GM), and added another enhancing tissue (ENH) class. STI requires tissue masks to compute joint histograms. To compute masks for WM+GM class, we combined WM and GM probabilistic tissue priors from the MNI152 atlas available via the FSL package^23^, thresholded them at 80% and resampled to Tal111 space. ENH masks were obtained by applying brain mask and thresholding. We labeled this normalization method STI-c to signify that it is designed specifically for contrast enhanced MRI. Figure 3 illustrates the effect of both normalization approaches on the resulting image histograms.

**Figure 3.**
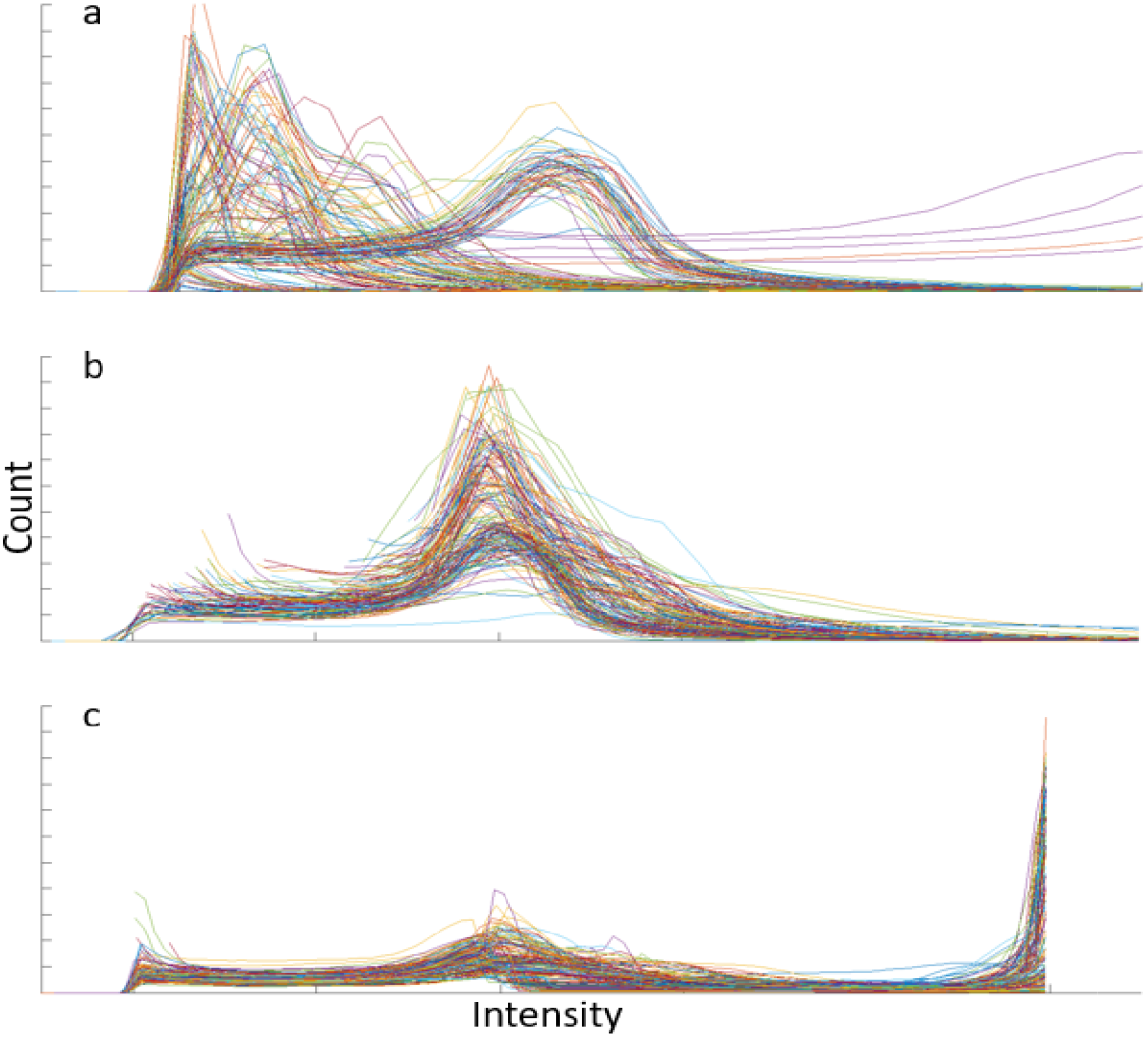
Effects of normalization on histogram of 160 T1c images. (a) histogram of non-normalized data, (b) linear z transform, (c) STI-c normalization.

**Figure 4.**
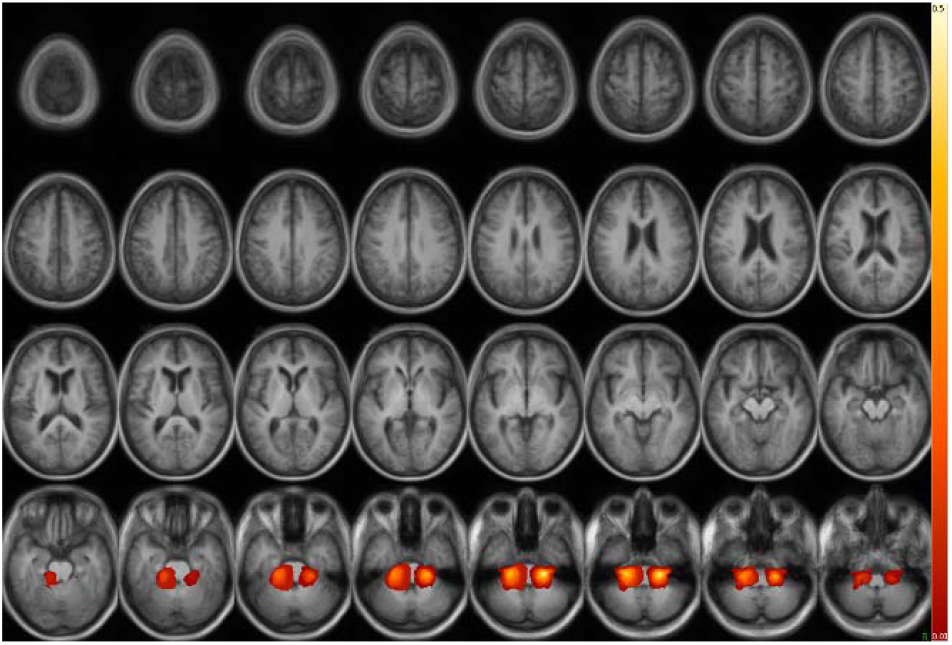
VS pseudo-probabilistic atlas of VS in a standard brain space. The atlas is derived from the 29 GKRS plans (pFOV-29

### 2.3. Segmentation of schwannoma on T1c images

#### 2.3.1. CNN model architecture and inputs

To segment schwannoma in T1c images, we adapted the DeepMedic convolutional neural network (CNN) architecture^24^. This 3D architecture has shown good performance on the task of glioma segmentation. It implements receptive field of 17^3^ voxels, equivalent to 17^3^ mm in Tal111 space, and supports multi-scale processing via parallel convolutional pathways, where the same source image can be input several times at different resolutions to make the classification aware of both local and global contextual features. DeepMedic targets to maximize the Dice coefficient between ground truth and predicted segmentations.

As the original DeepMedic CNN outputs five tissue classes and we were only interested in two (lesion/non-lesion), we reduced the number of computed features (equivalently, layer sizes) to both reduce the dimensionality of the CNN approximator and speed up training. Specific DeepMedic layer configuration parameters are shown in Table 1 of the Appendix. To implement multi-scale context awareness, we also enabled a second CNN pathway with input downsampled by the factor of two and receptive field of 34^3^ mm. This customized DeepMedic CNN architecture is further referred to as DM-s.

**Table 1.** Imaging features computed for each segmentation.

| Geometric features | Description/formula |
| --- | --- |
| Volume, mm<sup>3</sup> | Number of voxels × voxel volume |
| Compactness | (surface of sphere with the same volume)/(surface area) |
| Eccentricity | Long diameter/short diameter |
| Histogram features |  |
| Median | Median intensity |
| Standard deviation | Standard deviation of intensity |
| Robust minimum | 5<sup>th</sup> intensity percentile |
| Robust maximum | 95<sup>th</sup> intensity percentile |
| Texture homogeneity (1mm) | Sum(Pij/(1+(i-j)<sup>2</sup>)) where Pij are GLCM elements |

For DM-s training and inference, T1c images are resampled to Tal111 space and intensity-normalized using piecewise linear or STI-c method. Our final choice of specific normalization method will be discussed below. To supply anatomical priors to DM-s, we created an average anatomical atlas of lesion locations using lesion masks of subjects that contributed to the pFOV-29 template. These 29 lesion masks were resampled to Tal111 space and averaged. The resulting average image, referred to as pFOV-29-les (Fiugre 4), was then binarized and dilated using spherical kernel with 5mm radius. During the first pass of training and inference, the resulting bilateral mask was supplied to DM-s to confine training samples to the anatomically relevant region.

#### 2.3.2. Segmentation post-processing

To reduce the false positive segmentation labels common to CNN output and to improve the structure of resulting segmentations, various post-processing methods such as fully connected continuous random field (CRF)^24^ are used. However, when we tried to use the CRF post-processing on our data, per-case parameter tuning was required. Since schwannoma is more localized in the brain compared to glioma, we implemented our own procedure that uses anatomical priors to better localize the CNN output. Namely, we first extract all 1-connected components (CCs) from the DM-s output using 6-point 3D neighborhood for connectivity check. We then multiply the CCs voxel wise by pFOV-29 atlas mask. The resulting mask is thus weighted by prior probability of lesion appearance. Then for each CC we compute the voxel-wise sum of these weights, which may be interpreted as the cumulative probability of the CC to contain the actual lesion. The CC with maximum cumulative probability is then chosen as the final segmentation. This procedure is designed to eliminate false positive label outside of common schwannoma locations. As the final post-processing segmentation step, we apply two iterations of hole removal^25^ which smoothens mask topology without changing overall shape.

#### 2.3.3. DM-s training experiments

For training and validation experiments, out of 158 T1c images that were correctly coregistered to Tal111 space, we used 95 T1c images that had GK SRS planning segmentations confirmed by a neurosurgeon and a radiation oncologist available to serve as ground truth. This subset is further referred to as gt-95. The remaining subset of 63 images (nogt-63) without GK SRS segmentation mask, was used for manual validation, as detailed further.

For the first training experiment, we split the gt-95 into 80 training and 15 validation cases. T1c images were normalized with STI-c; for the input ROI, we used the original pFOV-29 ROI mask dilated with 5 mm kernel. Training was done with default settings (50 cases/1000 image patches loaded per epoch, batch size of 10) for 500 epochs. Training converged on test data with average testing Dice coefficient of 0.99, and average validation Dice coefficient of 0.79 that stabilized after 300 epochs. For the second experiment, we changed normalization to linear, leaving all other parameters unchanged. Similarly to the first experiment, Dice coefficient of 0.99 on training data was achieved after 500 epochs, but average validation Dice score improved to 0.89. As linear normalization appeared superior for this task, we excluded STI-c from all further experiments. For the third experiment, we reproduced the same setup but changed validation set to 15 different cases. The results were similar to previous experiment, confirming the robustness of the trained classifier. Finally, to improve generalizability, in the fourth experiment we retrained the model with the same parameters using all 95 cases from gt-95 as training set.

To test the performance on unseen data without ground truth, we applied the trained model from the fourth experiment, designated as DM-s4, to the nogt-63 dataset. To improve the outcomes, we applied the segmentation post-processing. Expert review of the segmentation results revealed 41 good segmentations, 5 where a small part of cyst (CST) or enhancement (ENH) was missed, and 17 cases where more than 20% of the tumor was misclassified, designated as failed. In the cohort of the failed cases, we identified three large groups: one where CST was missed, the other where lesion was outside the input ROI, and the third where tumor had an unusually complex composition and shape, often with multiple necrotic cores or displaced by ventriculomegaly (Figure 5).

**Figure 5.**
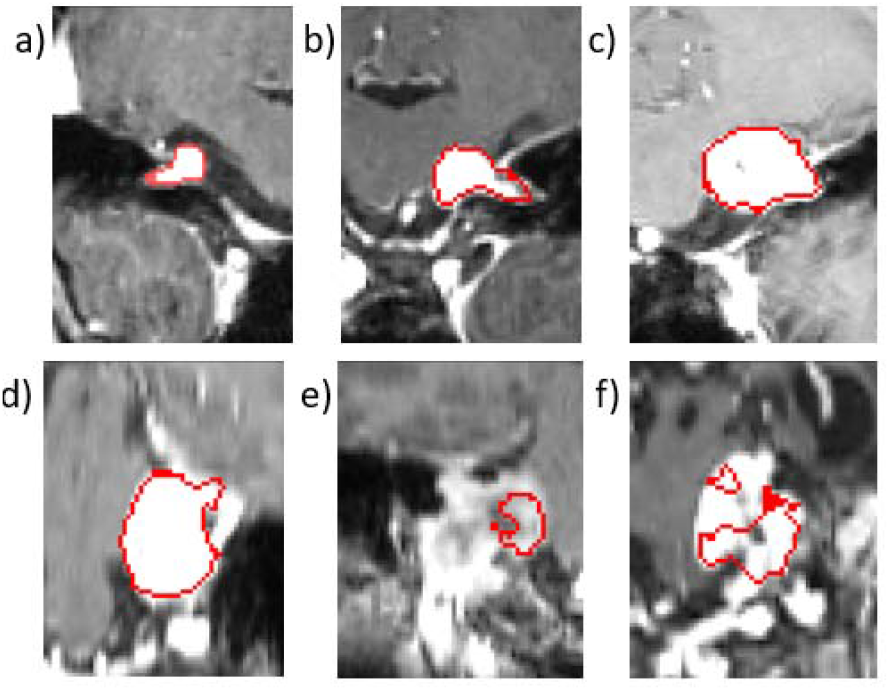
Typical good (a-c) and failed (d-f) segmentations by DM-s5.

To improve the coverage of the segmentation algorithm, in the fifth experiment for the input mask we used a 20 mm dilation instead of 5 mm. We also re-balanced the training set by increasing by 50% the frequency of cases with CST presented to CNN. The remaining training parameters and post-processing were unchanged from the previous experiment. Dice scores for each training epoch is shown on Figure 6. The final Dice scores for testing and validation sets were 0.99 and 0.92, accordingly. The newly trained model, designated as DM-s5, was tested on the nogt-63 dataset. Review identified 51 good segmentations, 4 acceptable and eight failed cases. Out of failed cases, four had more complex structure as shown in Figure DM-S-EXAMPLES. This final model was used for all further imaging feature evaluations, as detailed below.

**Figure 6.**
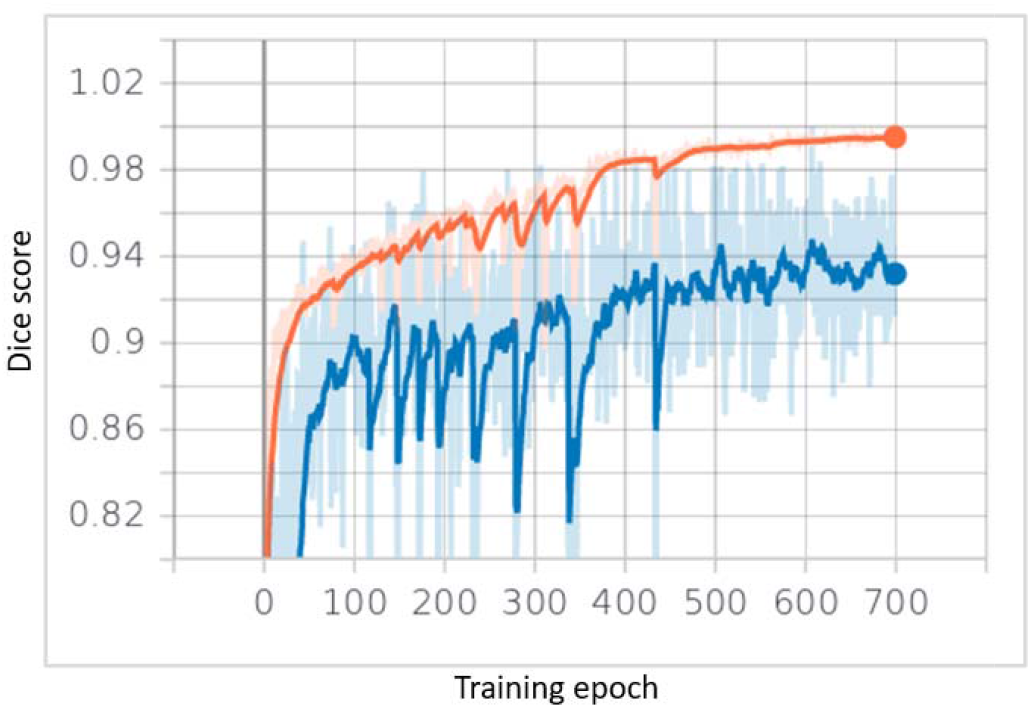
Training Dice scores for testing (orange) and validation (blue) sets during DM-s5 training.

#### 2.3.3. Validation by expert segmentation

For all expert segmentations, we used contouring feature of the online XNAT-OHIF viewer^16^. All lesion masks were resampled to Tal111 space using nearest neighbor interpolation. For comparison, we used Sorensen-Dice coefficient and average surface distance (ASD)^26^, also sometimes referred to as surface-to-surface distance (S2S), a measure that estimates accuracy of match of linear boundary contours rather than volumetric match. We also used a signed measure called ‘diameter difference’, that estimates the difference in linear dimension of two segmentations, by comparing the radii of spheres with volume equal to segmentation volume.

### 2.4. Segmentation based feature analysis

Based on T1c segmentation of VC, we examined five first order histogram features, as well as three geometric features (Table 1). These features were selected either because they are easy to interpret (e.g. median or standard deviation), or because they were previously reported to predict outcomes for VS (volume, homogeneity)^8,9^ or other tumor types^27^. The number of features was kept below 10 to reduce the likelihood of false discovery.

We then proceeded to evaluate feature robustness between segmentations. To this end, we compared features based on our own segmentations, DM-s5, and pre-GK SRS plans, with paired t-test. Pearson’s R correlation was also computed for the same sets.

### 2.5. Clinical outcome analysis

For a subset of participants from the original cohort of 158 images, we reviewed follow-up imaging radiology reports available via the hospital information system. Outcomes were initially defined as (a) VS growth and (b) VS stability and (c) VS shrinkage as determined by radiologist from each of the follow-up scans. Tumor growth was determined by a radiologist at the time of follow-up review. It was defined as at least 10% increase in the linear dimension of VS outline on the largest slice containing the VS. Outcomes were available for 64 cases from the gt-95 set and for 29 cases from the nogt-63 set. Tumor growth was recorded in 9 cases, shrinkage in 32, and stability in 23 cases for gt-95 set, and in the nogt-63 set 3 cases had growth, 15 cases demonstrated shrinkage and 11 were stable across follow-up. For the outcomes analyses, we therefore considered two types of non-response to GK SRS surgery: (a) VS growth and (b) VS non-decrease (growth or stability). The “non-decrease” outcome was considered solely to improve the statistical power of the outcome comparison, as decrease (growth) event was relatively rare. For each segmentation method and each feature, we computed Kaplan-Meier (KM) curves for these two outcomes, split by an optimized threshold into upper and lower curves. The threshold was selected by uniform sampling between 40^th^ and 60^th^ percentile of the cases by minimizing the p value of the log rank test for significant difference between the upper and lower KM curves. It should be noted that for this analysis, no follow-up images were analyzed, so the only source of outcome information were radiology reports.

## 3. Results

### 3.1. Inter-rater agreement

To evaluate the accuracy of segmentation, we first evaluated human expert agreement in segmenting VS on T1c images. To that end, two neurosurgery experts, (raters 1 and 2, HS and KV), segmented ten lesions selected randomly from the images with ground truth (gt-95 set), and the results were compared to the ground truth from the GK SRS planning workstation. The results are shown in Table 2.

**Table 2.** Agreement of manual segmentations averaged over 10 cases from the gt-95 set. Diameter difference is between rater in column and rater in row.

| Rater | Dice coefficient |  |  | ASD, mm |  |  | Diameter difference, % |  |  |
| --- | --- | --- | --- | --- | --- | --- | --- | --- | --- |
|  | Rater 1<br>(HS) | Rater 2<br>(KV) | GK SRS<br>plan | Rater 1<br>(HS) | Rater 2<br>(KV) | GK SRS<br>plan | Rater 1<br>(HS) | Rater 2<br>(KV) | GK SRS<br>plan |
| Rater 1<br>(HS) | 1 | 0.83 | 0.91 | 0 | 0.6 | 0.35 | 0 | 7.0 | 3.3 |
| Rater 2<br>(KV) | 0.83 | 1 | 0.89 | 0.6 | 0 | 0.37 | -7.0 | 0 | 3.9 |

**Table 3.** Agreement of Rater 3, GK planning and DeepMedic (DM-s5) segmentations.

| Rater | Dice coefficient |  |  | ASD, mm |  |  | Diameter difference, % |  |  |
| --- | --- | --- | --- | --- | --- | --- | --- | --- | --- |
|  | Rater 3 (KC) | GK SRS plan | DM-s5 | Rater 3 (KC) | GK SRS plan | DM-s5 | Rater 3 (KC) | GK SRS plan | DM-s5 |
| Rater 3 (KC) | 1 | 0.85 | 0.79 | 0 | 0.7 | 1.05 | 0 | -7.2 | -10.8 |
| GK SRS plan | 0.85 | 1 | - | 0.7 | 0 | - | 7.2 | 0 | - |

### 3.2. Agreement between an expert and DM-s

Another neurosurgery expert (Rater 3, KC) segmented all available T1c images, and segmentations were compared to gt-95 dataset and to those in nogt-63 without ground truth segmented by DM-s5 model. Eight cases failed by DM-s5 were excluded from comparison. Comparison results are shown in Table 2. We did not compare our DM-s5 model to segmentations from gt-95 since the former used the latter as ground truth for training with 99% Dice score achieved on the training set (Figure 6).

### 3.3 Feature robustness

Results of feature wise comparisons across different segmentation methods are shown in Table 4. Comparison between different human raters (Rater 3 and GK SRS plan) revealed that only eccentricity and sphericity averages were preserved across raters. Both features were also robust across human and AI segmentations with mean absolute difference below 3%. Homogeneity on average was the most stable across segmentation methods with mean absolute difference below 2% for all cases. Also, notably, maximum intensity and volume means were significantly different across segmentation methods but had perfect correlation.

**Table 4.** Robustness metrics for each feature between different segmentation methods. P values are shown for the null hypothesis that feature means are not different according to paired two-tailed t test.

| Feature | Rater 3/GK SRS plan |  |  | Rater 3/DM-s5 |  |  |
| --- | --- | --- | --- | --- | --- | --- |
|  | p value | Mean absolute difference, % | Pearson's R | p value | Mean absolute difference, % | Pearson's R |
| Homogeneity | 0.01 | 1.9 | 0.95 | 5.32e-03 | 0.2 | 0.90 |
| Min | 3.37e-24 | 142 | 0.72 | 5.92e-18 | 179 | 0.55 |
| Median | 2.81e-15 | 16 | 0.91 | 1.41e-13 | 56 | 0.82 |
| Max | 9.65e-14 | 1.5 | 1.00 | 4.70e-13 | 15 | 1.00 |
| Std | 1.50e-25 | 16 | 0.89 | 1.12e-17 | 16 | 0.92 |
| Volume | 4.22e-14 | 16 | 0.99 | 1.62e-08 | 33 | 0.98 |
| Eccentricity | 0.1 | 1.9 | 0.90 | 0.05 | 3.4 | 0.86 |
| Sphericity | 0.61 | 0.6 | 0.95 | 2.42e-04 | 2.7 | 0.90 |

### 3.4. Robustness of clinical outcome analysis

The p values of logrank test for the difference between upper and lower curves in Kaplan-Meier analysis, as well as optimized split threshold percentiles, are shown in Table 5 for growth outcome and in Table 6 for no-shrinkage outcome.

**Table 5.**
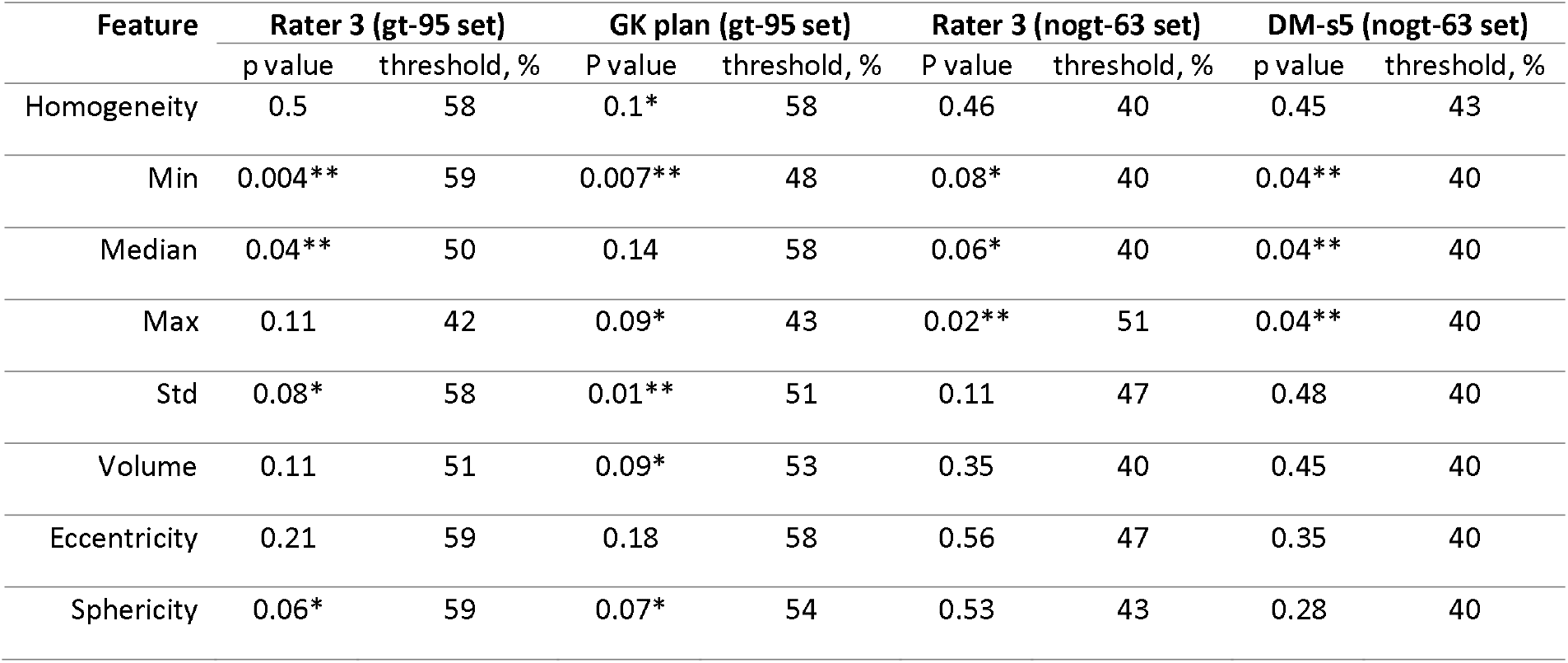
Logrank test p values and optimized threshold for Kaplan-Meier curves defined for VS growth outcome, using different segmentation methods for each examined feature, for gt-95 and nogt-63 datasets.

| Feature | Rater 3 (gt-95 set) |  | GK plan (gt-95 set) |  | Rater 3 (nugt-63 set) |  | DM-s5 (nugt-63 set) |  |
| --- | --- | --- | --- | --- | --- | --- | --- | --- |
|  | p value | threshold, % | P value | threshold, % | P value | threshold, % | p value | threshold, % |
| Homogeneity | 0.5 | 58 | 0.1* | 58 | 0.46 | 40 | 0.45 | 43 |
| Min | 0.004** | 59 | 0.007** | 48 | 0.08* | 40 | 0.04** | 40 |
| Median | 0.04** | 50 | 0.14 | 58 | 0.06* | 40 | 0.04** | 40 |
| Max | 0.11 | 42 | 0.09* | 43 | 0.02** | 51 | 0.04** | 40 |
| Std | 0.08* | 58 | 0.01** | 51 | 0.11 | 47 | 0.48 | 40 |
| Volume | 0.11 | 51 | 0.09* | 53 | 0.35 | 40 | 0.45 | 40 |
| Eccentricity | 0.21 | 59 | 0.18 | 58 | 0.56 | 47 | 0.35 | 40 |
| Sphericity | 0.06* | 59 | 0.07* | 54 | 0.53 | 43 | 0.28 | 40 |

**Table 6.** Logrank test p values and optimized threshold for Kaplan-Meier curves defined for VS non-shrinkage outcome, using different segmentation methods for each examined feature, for gt-95 and nogt-63 datasets.

| Feature | Rater 3 (gt-95 set) |  | GK plan (gt-95 set) |  | Rater 3 (nugt-63 set) |  | DM-s5 (nugt-63 set) |  |
| --- | --- | --- | --- | --- | --- | --- | --- | --- |
|  | p value | threshold, % | P value | threshold, % | P value | threshold, % | p value | threshold, % |
| Homogeneity | 0.19 | 40 | 0.59 | 51 | 0.21 | 40 | 0.16 | 51 |
| Min | 0.11 | 42 | 0.21 | 42 | 0.06* | 40 | 0.05** | 51 |
| Median | 0.54 | 40 | 0.25 | 58 | 0.5 | 43 | 0.33 | 40 |
| Max | 0.35 | 42 | 0.16 | 50 | 0.3 | 58 | 0.13 | 43 |
| Std | 0.52 | 50 | 0.19 | 47 | 0.25 | 58 | 0.22 | 47 |
| Volume | 0.34 | 42 | 0.38 | 59 | 0.31 | 58 | 0.06* | 47 |
| Eccentricity | 0.07* | 48 | 0.13 | 45 | 0.08* | 51 | 0.11 | 54 |
| Sphericity | 0.45 | 53 | 0.38 | 40 | 0.12 | 58 | 0.32 | 40 |

For predicting growth, minimum, median and maximum were either significant or trending for both manual and DM-s5 segmentation methods for both examined datasets. Sphericity was trending for gt-95 dataset for both manual methods. No significance was found for other features for nogt-63 dataset, likely due to low number of recorded growth events (3). Volume had some predictive power in the larger gt-95 dataset, but not in the nogt-63.

For predicting non-shrinkage, eccentricity was trending for all methods. Min was trending for both methods only for nogt-63 dataset.

### 3.5. Implementation

MGA prerpocessing, as well as DeepMedic training, validation and testing was performed on a single core CPU with GeForce RTX 2080 Ti GPU. Training took approximately 3 hours per experiment, whereas pre- and post-processing took under 1 minute per subject image. Feature calculations were implemented using in-house shell scripts and C++ libraries. Correlation and t-test analyses were performed using Python library scipy v. 1.3.1 and Kaplan-Meier analyses using Python library lifelines v. 0.26.4. All Python analyses were performed in Jupyter notebooks environment.

The pipeline using MGA preprocessing and DeepMedic segmentation, was implemented as a Docker image, available at https://github.com/satrajitgithub/mga_schwannoma. This image is integrated with XNAT container service [https://bitbucket.org/xnatdev/container-service] and XNAT/OHIF viewer^16^, allowing to produce segmentations on data archived in XNAT, and also modify and store them with the original archived DICOM MR images.

### 3.6. Data Availability

Data has been made available in the Cancer Imaging Archives^28^ under project name CONDR-Vestibular Schwannoma. This dataset includes 229 unprocessed DICOM scans, 158 MGA co-registered DICOM scans, 95 GK SRS ground truth segmentations, and 178 expert manual segmentations (Rater1=10, Rater2=10, Rater3=158). Radiomic measures are available for all 158 sessions and outcome measures are provided for 93 patients with follow-up imaging. In addition, the CONDR-Vestibular Schwannoma dataset on TCIA will include unprocessed follow-up scans. All data in TCIA has undergone standard de-identification procedures and is available under a variant of Creative Commons license.

## 4. Discussion

In this work, we present a framework of methods to analyze (VS) from T1-weighed post-contrast MRI (T1c). Building on previous work, we developed spatial and intensity normalization preprocessing. We further built a probabilistic atlas of VS from 26 cases. Based on these preprocessing steps, we developed and trained a CNN model labeled DM-s5 to autonomously segment VS on T1c images. Segmentations from GK SRS plan (gt-95 dataset) were used as ground truth. We then compared VS segmentations between the GK SRS plan, DM-s5 output and segmentations performed by an experienced neurosurgery resident. The resulting model performed exceptionally on gt-95 with Dice score of 0.99 for training and 0.92 for validation subsets, on par with performance reported by other studies^11,12^. However, performance was lower on the additional validation set (nogt-63) where ground truth was established by a different rater (Rater 3) and using different segmentation software (XNAT/OHIF viewer). This supports the notion that CNN segmentations produced by models informed by different initial labeling methods and raters may not be fully comparable or interoperable.

In our subsequent evaluations, we investigated the robustness of select radiomic features to different segmentation methods and established the most robust features. We concluded that systematic biases that exist between manual raters, were also learned by the DM-s5 algorithm (Table 3). Therefore, if a pre-trained AI tool were to be shipped to clinic, detailed clinical setting and provenance of the ground truth dataset must be carefully evaluated to be in line with the use of the AI segmentation tool. In our case, GK SRS plans, having the primary purpose of outlining the irradiation region, tended to over-estimate the VS area. Another bias in the GK planning is tendency to decrease dose to facial nerve, which is anterior and superior in the tumor. This leads to decreased coverage to this area of the tumor. On the other hand, Rater 3 aimed to maximize anatomical accuracy, which resulted in systematic bias in estimated volume between these two methods.

Our work also investigated the major issue of stability of radiomic features between different segmentation methods as applied to VS on T1c images. We found that all features except sphericity and eccentricity, do not preserve their means when different segmentation methods are used. However, for

T1c images, volume and robust maximum intensity correlated perfectly between all segmentation methods. Another class of features included those which correlated relatively well (Pearson’s R over 0.9) and had a low (below 3%) mean absolute difference. Those included homogeneity, eccentricity and sphericity.

Finally, we investigated the effect of different segmentation methods on the clinical outcomes analysis, by comparing the Kaplan-Meier feature-split curves for each radiomic feature based on different segmentation methods. We found that the robust maximum and minimum intensities were the most consistently predictive of VS growth across all segmentation methods and subsets of data (Table 5). Robust minimum, however, was also the least stable across segmentation methods, which makes it poor choice for clinical outcomes analysis. We also found that segmentation method did not significantly affect the shape of KM curves (Figure 7). No statistical significance detected in the difference between upper and lower curves for nogt-63 dataset may be explainable by the low number of recorded growth events (3 out of 29). We also investigated which features predicted that VS would not shrink post-GK SRS, with elongated tumors (high eccentricity) found to be less susceptible to contraction (Figure 8).

**Figure 7.**
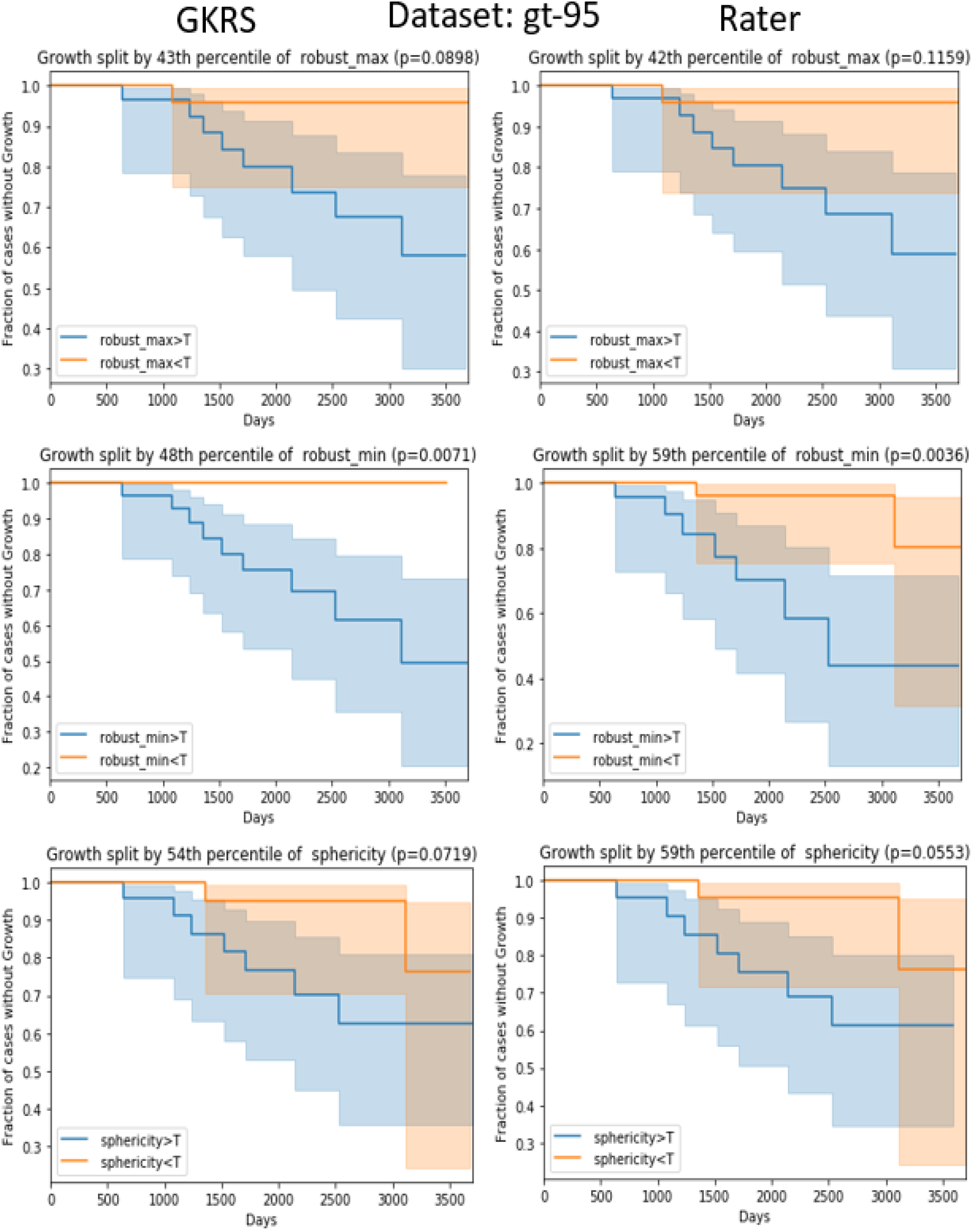
Kaplan-Meier curves for VS growth event for gt-95 dataset based on GKRS plan (left column) and Rater 3 segmentations (right column) split by robust maximum (top row), minimum (middle row), and sphericity (bottom row). Orange and blue transparent areas show the 95% confidence ranges for upper and lower curves.

**Figure 8.**
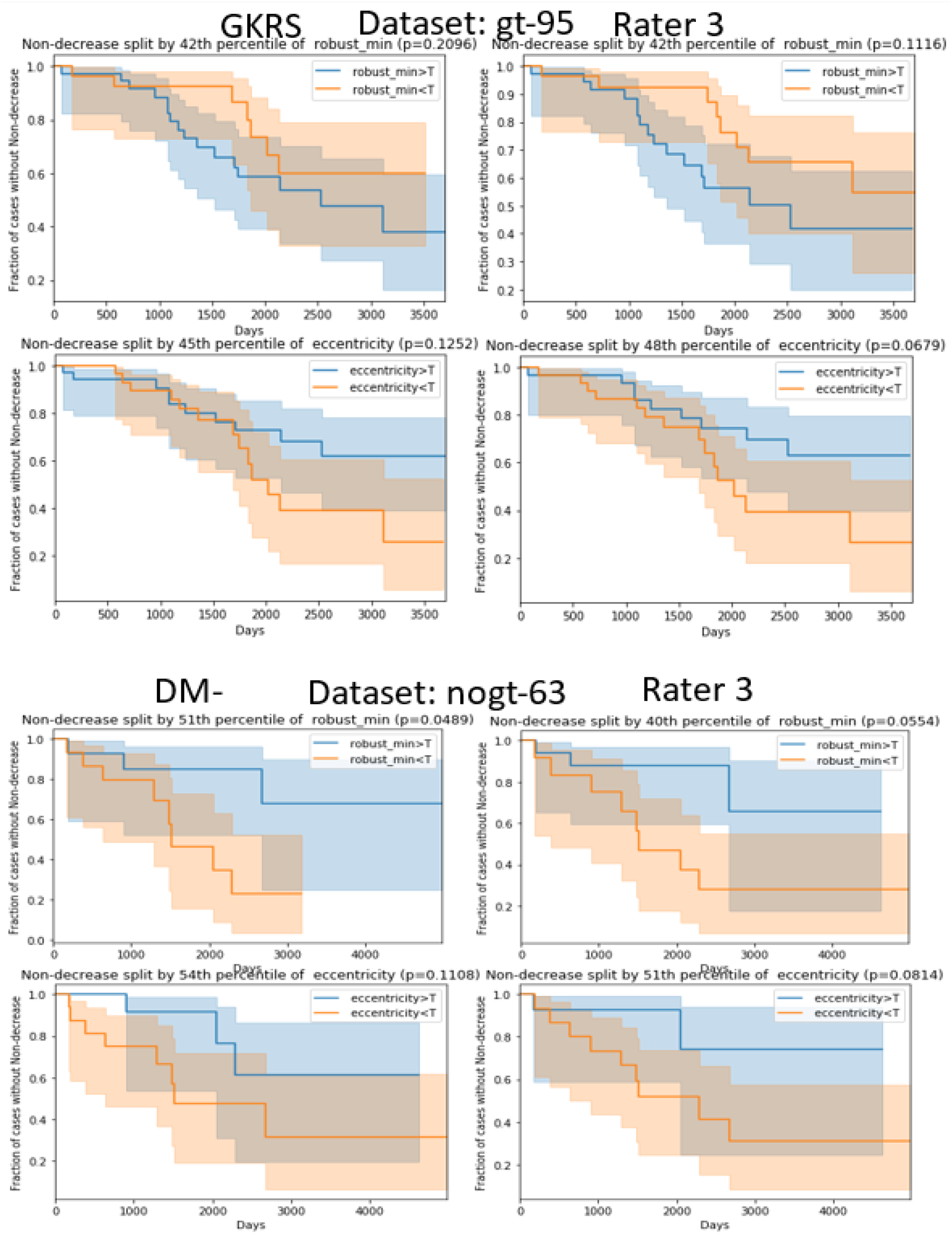
Kaplan-Meier curves for VS non-shrinkage event for robust minimum and eccentricity acquired from segmentations by Rater 3 and GKRS plan (gt-95 dataset) and from segmentations by DM-s5 and Rater 3 (nogt-63 dataset). Orange and blue transparent areas show the 95% confidence ranges for upper and lower curves.

Compared to other studies^12^, our CNN segmentation method DM-s5 uses only T1c images, which makes it both more practical and potentially less accurate. Lower performance on nogt-63 dataset can be explained by the difference in manual segmentation methods used to produce ground truth segmentations. In addition, partial brain field of view introduced another source of variability that might have affected the performance, since DM-s5 uses whole brain area in its dual resolution input to increase the context sensitivity.

Overall, this manuscript describes one of the first attempts to develop a united automated framework for segmentation and radiomic analysis of VS using T1c images. This effort is also unique in evaluating the effect of various segmentation methods and radiomic feature selection on the robustness of outcomes analysis in VS.

In the future, we plan to validate our findings on external datasets and with other segmentation tools on our data. To facilitate that, in addition to releasing the VS processing pipeline for research use, we are releasing our T1c sets and ground truth segmentations in the TCIA^28^.

## Data Availability

All data produced in the present study are available upon reasonable request to the authors

## Abbreviations

VS: Vestibular Schwannoma
MGA: Multimodal Glioma Analysis
GK: Gamma Knife
QC: Quality Control
NCR: necrosis
ENH: enhancement
ROI: region of interest
FOV: field of view
nogt-63: no gamma knife contour
CNN: convolutional neural network
gt: gamma knife
DM: DeepMedic

## Conflicts of Interest

AHK is a consultant for Monteris Medical.

## Notes

### Competing Interest Statement

Albert Kim is a consultant for Monteris Medical.

### Funding Statement

This work was partially funded by NCI grant
U24CA204854 aimed at developing data and knowledge management platforms to support the collection, analysis, and sharing of imaging and related data for use in cancer research.

### Author Declarations

IRB of Washington University in Saint Louis gave ethical approval for this work.

